# Biallelic and *de novo* variants in *ATP6V0A1* cause progressive myoclonus epilepsy and developmental and epileptic encephalopathy

**DOI:** 10.1101/2021.06.01.21257500

**Authors:** Laura C Bott, Mitra Forouhan, Maria Lieto, Ambre J Sala, Ruth Ellerington, Janel O Johnson, Alfina A Speciale, Chiara Criscuolo, Alessandro Filla, David Chitayat, Andrea H Nemeth, Italian Undiagnosed Diseases Network, Francesco Angelucci, Wooi Fang Lim, Pasquale Striano, Federico Zara, Ingo Helbig, Mikko Muona, Carolina Courage, Anna-Elina Lehesjoki, Samuel F Berkovic, V-ATPase Consortium, Kenneth H Fischbeck, Francesco Brancati, Richard I Morimoto, Matthew JA Wood, Carlo Rinaldi

**Affiliations:** Department of Molecular Biosciences, Rice Institute for Biomedical Research, Northwestern University, Evanston, Illinois 60208, USA; Department of Paediatrics, University of Oxford, Oxford OX1 3QX, UK; Department of Physiology, Anatomy and Genetics, Oxford OX1 3QX, UK; Department of Neurosciences, Reproductive and Odontostomatological Sciences Federico II University, Naples 80121, Italy; Neuromuscular Diseases Research Section, Laboratory of Neurogenetics, National Institute on Aging, National Institutes of Health, Bethesda, Maryland 20892, USA; Division of Clinical and Metabolic Genetics, Department of Pediatrics, The Hospital for Sick Children, University of Toronto, Toronto, Ontario M5G 1X8, Canada; The Prenatal Diagnosis and Medical Genetics Program, Department of Obstetrics and Gynecology, Mount Sinai Hospital, University of Toronto, Toronto, Ontario M5G 1X5, Canada; Nuffield Department of Clinical Neurosciences, University of Oxford, Oxford OX3 9DU, UK; Department of Life, Health and Environmental Sciences, University of L’Aquila, 67100 Coppito, L’Aquila, Italy; Institute for Research, Hospitalization and Health Care (IRCCS) “G. Gaslini” Institute, Genova 16147, Italy; Division of Neurology, Children’s Hospital of Philadelphia; The Epilepsy NeuroGenetics Initiative (ENGIN), Children’s Hospital of Philadelphia; Department of Biomedical and Health Informatics (DBHi), Children’s Hospital of Philadelphia; Department of Neurology, University of Pennsylvania, Perelman School of Medicine, Philadelphia, Pennsylvania 19104, USA; Blueprint Genetics, 02150 Espoo, Finland; Folkhälsan Research Center, Helsinki, Finland; Department of Medical and Clinical Genetics, Medicum, University of Helsinki, Helsinki 00290, Finland; Epilepsy Research Centre, Department of Medicine, University of Melbourne, Austin Health, Heidelberg, Victoria 3010, Australia; Neurogenetics Branch, National Institute of Neurological Disorders and Stroke, National Institutes of Health, Maryland 20892, USA; IRCCS San Raffaele Pisana, 00163 Roma, Italy; Oxford Harrington Rare Disease Centre, University of Oxford, Oxford OX1 3QX, UK

**Keywords:** V-ATPase, epileptic encephalopathy, *C. elegans* disease modelling, organelle acidification, lysosomal disease

## Abstract

The vacuolar H^+^-ATPase is a large multi-subunit proton pump, composed of an integral membrane V0 domain, involved in proton translocation, and a peripheral V1 domain, catalysing ATP hydrolysis. This complex is widely distributed on the membrane of various subcellular organelles, such as endosomes and lysosomes, and plays a critical role in cellular processes ranging from autophagy to protein trafficking and endocytosis. Variants in *ATP6V0A1*, the brain-enriched isoform in the V0 domain, have been recently associated with developmental delay and epilepsy in four individuals. Here we identified 17 individuals from 14 unrelated families with both with new and previously characterised variants in this gene, representing the largest cohort to date. Five affected subjects with biallelic variants in this gene presented with a phenotype of early-onset progressive myoclonus epilepsy with ataxia, while 12 individuals carried *de novo* missense variants and showed severe developmental and epileptic encephalopathy. The R740Q mutation, which alone accounts for almost 50% of the mutations identified among our cases, leads to failure of lysosomal hydrolysis by directly impairing acidification of the endolysosomal compartment, causing autophagic dysfunction and severe developmental defect in *C. elegans*. Altogether, our findings further expand the neurological phenotype associated with variants in this gene and provide a direct link with endolysosomal acidification in the pathophysiology of *ATP6V0A1*-related conditions.

## Introduction

Vacuolar-type ATPases (V-ATPases) are a ubiquitous, multi-subunit, membrane-embedded rotary proton-pumping motor, arranged into a dissociable peripheral V1 domain (subunits A, B, C, D, E, F, G and H), responsible for ATP hydrolysis, and an integral membrane V0 domain (subunits a, c, c’, c”, d, e), responsible for proton translocation (Mazhab-Jafari et al., 2016; Roh et al., 2018; Zhao, Benlekbir, & Rubinstein, 2015). Through vesicular, luminal and extracellular acidification, the V-ATPases play a critical role in a number of both physiological and pathological cellular processes, from membrane trafficking and substrate degradation in lysosome, to viruses and bacterial toxins internalization and cancer growth and invasion (Vasanthakumar & Rubinstein, 2020). Other non-canonical roles of the V-ATPase complex, which are not readily attributable to its proton-moving activity, include membrane fusion, nutrient sensing, and scaffold for protein-protein interactions (Hiesinger et al., 2005; Hurtado-Lorenzo et al., 2006; Zhang et al., 2014). Recent studies have highlighted the critical importance of the V-ATPase complex in neuronal homeostasis and dysregulation of pH has been shown to be a converging pathogenic mechanism for several diseases, including Alzheimer’s (Avrahami et al., 2013; Lee et al., 2015), Parkinson’s (Betarbet et al., 2000; Boland et al., 2018; Dehay et al., 2010; Pal et al., 2016; Wallings, Connor-Robson, & Wade-Martins, 2019), amyotrophic lateral sclerosis (Şentürk, Mao, & Bellen, 2019; Yang & Klionsky, 2020), and lysosomal storage diseases (LSD) (Folts, Scott-Hewitt, Pröschel, Mayer-Pröschel, & Noble, 2016; Futerman & Van Meer, 2004). Mutations in subunits of the V-ATPase have been associated with autosomal recessive osteopetrosis (*ATP6V0A3*) (Frattini et al., 2000; Kornak et al., 2000) cutis laxa (*ATP6V0A2*) (Kornak et al., 2008), distal renal tubular acidosis (*ATP6V1B1, ATP6V0A4*) (Esmail et al., 2018; Karet et al., 1999; Smith et al., 2000), and epileptic encephalopathy (*ATP6V1A*) (Fassio et al., 2018), suggesting that this is an emerging class of human genetic disorders.

Some of the isoforms have different expression patterns in various tissues, which may allow the regulation of V-ATPase in a cell type and subcellular compartment specific manner. *ATP6V0A1* is the brain-enriched isoform of the a subunit in the V0 domain (Morel, 2003) and is part of the CLEAR (Coordinated Lysosomal Expression and Regulation) network of genes regulated by the master transcription factor EB (TFEB) (Palmieri et al., 2011).

Here we report 5 individuals from 2 unrelated families with compound heterozygous variants in *ATP6V0A1* presenting with a phenotype of early-onset progressive myoclonus epilepsy (PME) and ataxia and 12 cases with *de novo* missense variants in the same gene, with severe developmental and epileptic encephalopathy (DEE), resulting from direct impairment of endolysosome acidification and failure of lysosomal functions.

## Results

We have previously identified an Italian family with four members affected by early-onset progressive myoclonus epilepsy (PME) with ataxia and mental retardation, of unknown origin (Coppola et al., 2005). Seeking to unravel the genetic cause of this condition, we performed whole exome sequencing (WES) in 3 affected and 2 unaffected individuals (Fam. I; **Fig. 1A, Supplementary Fig. 1**) and found two previously unknown, compound heterozygous variants, c.445delG (p.E149Kfs18) and c.1483C>T (p.R495W) in *ATP6V0A1* (NM_001130021.3), that co-segregate with the clinical phenotype (**Fig. 1A**). From an independent WES screening of 85 PME patients, we identified 1 individual also from Italy with overlapping clinical features, harbouring the same compound heterozygous mutations (Fam. II.1; **Fig. 1A, Table 1**). Identity by descent analysis showed that the two families are unrelated (PI-HAT value = 0). While the c.445delG is not found in the gnomAD database (Lek et al., 2016), the allelic carrier frequency of the c.1483C>T variant (rs781278654) is 0.00001315 in the control population and predicted to be damaging (PolyPhen score: 0.99). Neither variant was present in an independent cohort of 200 Italian subjects, suggesting low frequency in this population.

**Figure 1:**
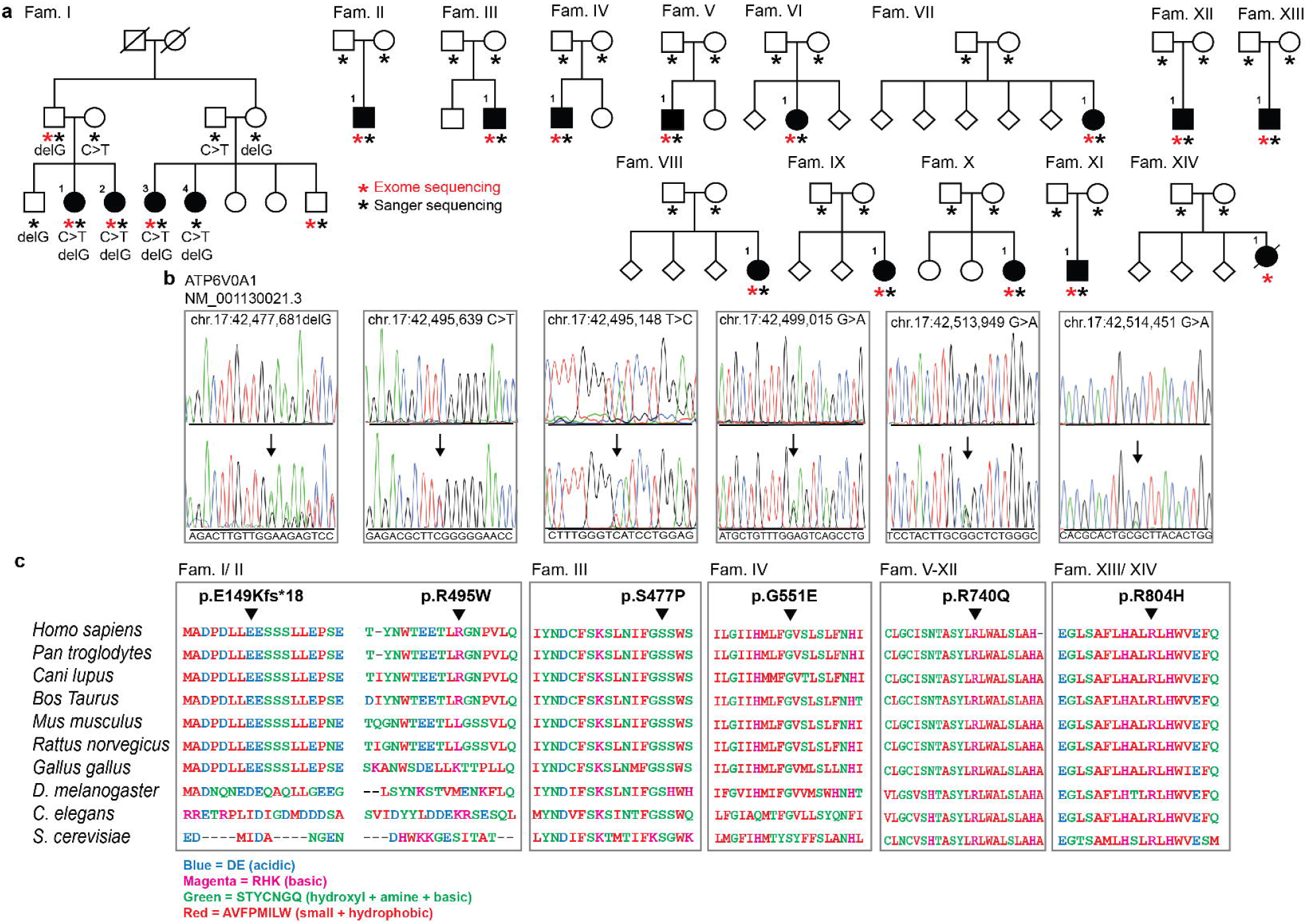
Identification of disease mutations in *ATP6V0A1*. (**A**) Pedigrees of all families included in the study are shown. Filled symbols represent affected individuals and are numbered within the family. Asterisks indicate subjects who underwent whole-exome sequencing (red) or confirmatory Sanger sequencing (black). Abbreviations: Fam. = family. (**B**) Representative electropherograms of genomic DNA sequencing of unaffected (top panel) and affected (bottom panel) individuals with identified mutations indicated by arrows. Gene symbol, reference sequence and genomic position of the changes are displayed above the electropherograms. (**C**) Sequence alignment of ATP6V0A1 protein across multiple species shows evolutionary conservation of the identified mutated residues, indicated by the black arrowhead, and surrounding regions. Gaps (black lines) are inserted between residues so that identical or similar amino acids are aligned in successive columns. Acidic residues (Asp, Glu) are in blue, basic residues (Arg, His, Lys) are in magenta, uncharged polar amino acids (Ser, Thr, Tyr, Asn, Gln) and Gly and Cys are in green, and nonpolar amino acids aside from Gly and Cys (Ala, Val, Phe, Pro, Met, Ile, Leu, Trp) are in red.

**Table 1.**
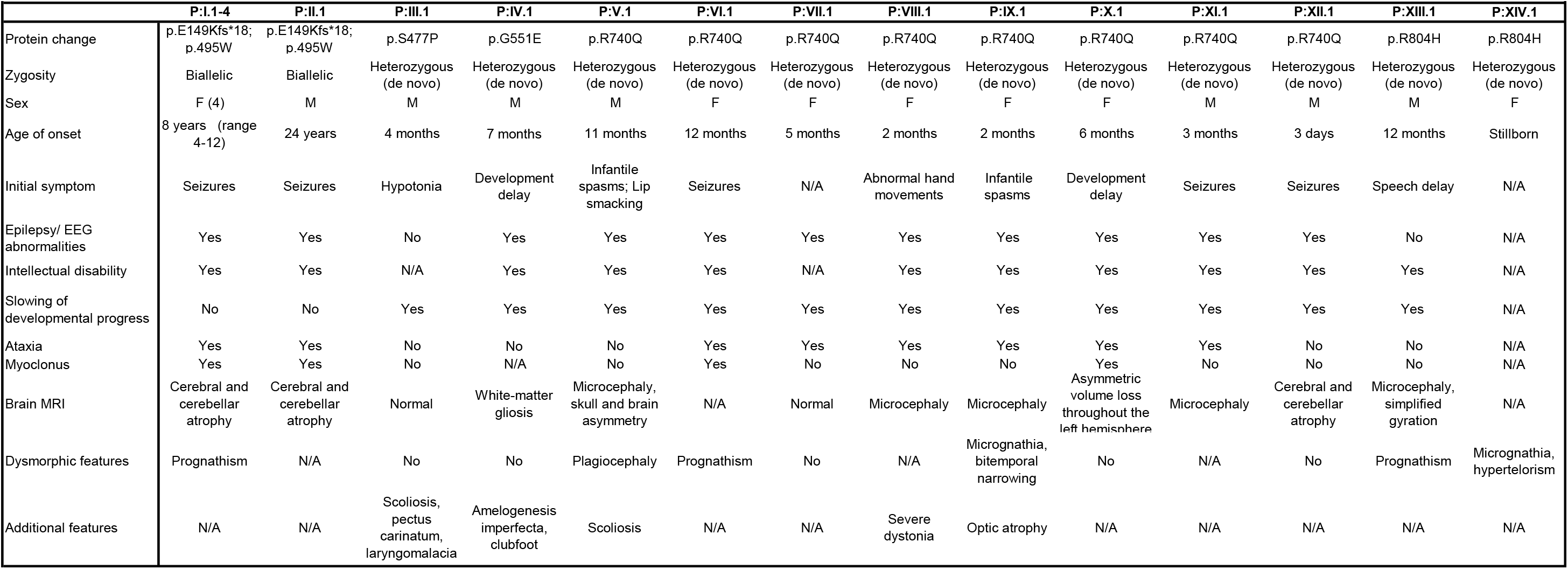
Clinical characteristics.

We next pooled genetic and clinical de-identified data of patients with severe developmental disorder (DD) enrolled in the Italian Undiagnosed Rare Diseases Network (IURDN) (n=110) (Salvatore et al., 2020) and the Deciphering Developmental Disorders study (n = 13,462) (Barash et al., 2010) and identified 3 individuals harbouring the *de novo* variants c.1652G>A (p.G551E) (Fam. IV.1) and c.2219G>A (p.R740Q) (Fam. V/VI.1) heterozygous variants in *ATP6V0A1* (**Fig. 1A**). Lastly we included in the study 9 additional individuals with unresolved DD who underwent diagnostic WES through GeneDx, Peking University First Hospital, and Pitié Salpêtrière University Hospitals, carrying the c.1429T>C (p.S477P) (Fam. III.1), c.2219G>A (p.R740Q) (Fam. VII-XII.1) and c.2411G>A (p.R804H) (Fam. XIII/XIV.1) *de novo ATP6V0A1* variants (**Fig. 1A**). We confirmed the presence of the mutations detected by WES in all individuals by Sanger sequencing and provided segregation data in immediate-relative carriers (**Fig. 1B**). The mutated residues in *ATP6V0A1* are evolutionarily conserved (**Fig. 1C**).

Altogether, we identified 17 affected subjects with mutations in *ATP6V0A1*, with p.R740Q being the most recurrent variant (8 subjects) (**Fig. 1A, Table 1**). The mean age of onset was 11.8 ± 7.5 years for individuals carrying the compound heterozygous mutations and 5.8 ± 4.2 months for individuals with the *de novo* mutations (DNMs) (**Table 1**). Subjects with the *de novo* variants showed a phenotype of developmental and epileptic encephalopathy (DEE), with frequent seizures, which were refractory to treatment, and severe mental retardation after a period of apparently normal development, with delay or loss of psychomotor milestones (**Table 1**). Several subjects were found to have microcephaly and other facial and skeletal dysmorphisms (**Table 1, Supplementary Fig. 2A and B**). Brain biopsy of a stillborn infant carrying the p.R804H variant showed swollen neurons and axons, with accumulation of granular, PAS-positive material, consistent with LSD with neuroaxonal dystrophy (**Fig. 1A, Fig. 2A**), while the placenta derived from the mother showed no evidence of storage cells (not shown).

**Figure 2:**
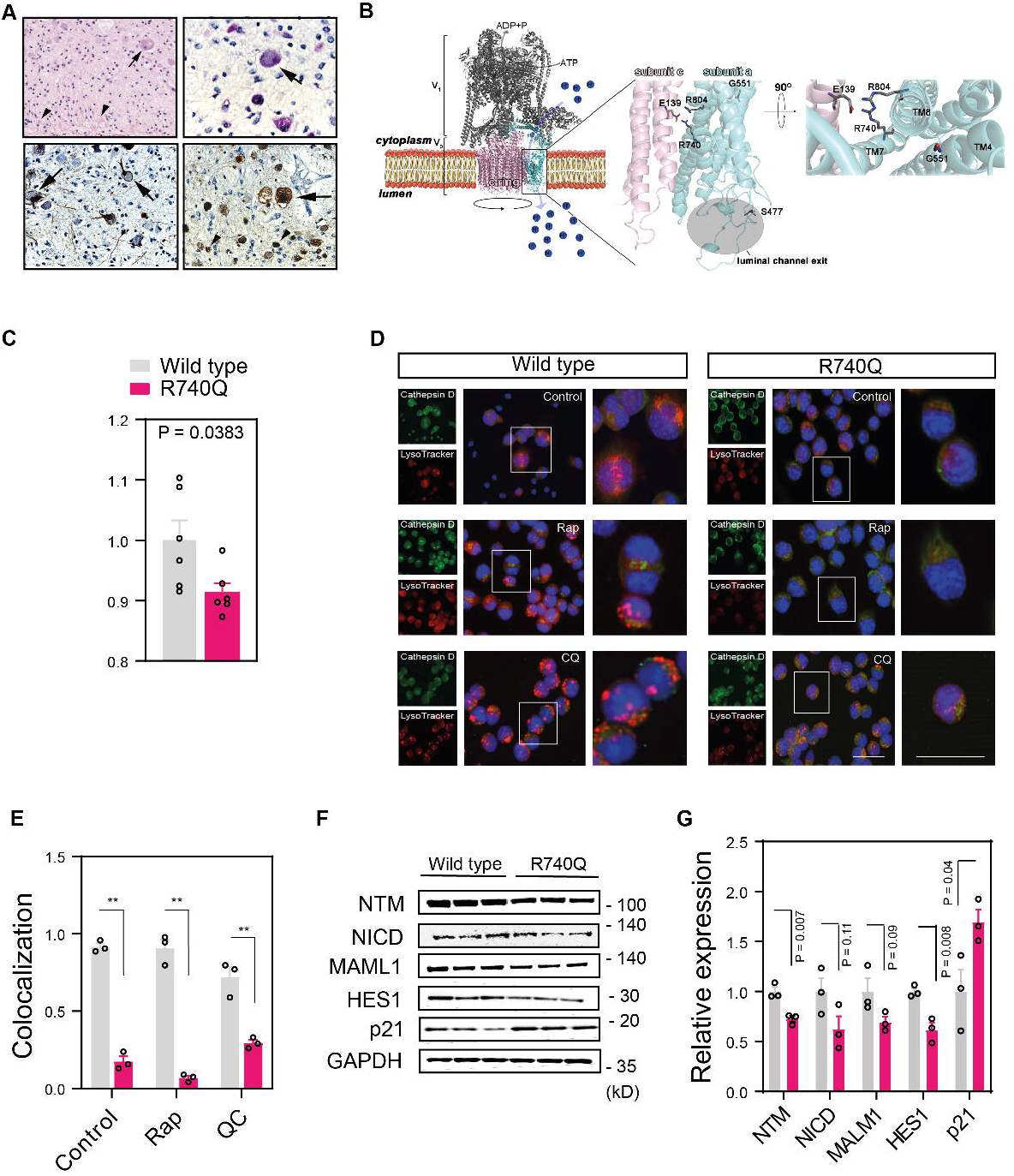
*De novo ATP6V0A1* mutations impair endolysosome acidification and activity. (**A**) Brain histopathology of subject P:XIV.1 shows swollen neurons (arrows) and axons (arrowheads) with granular periodic acid-Schiff (PAS)-positive deposits (top left panel: haematoxylin and eosin staining; top right panel: PAS staining; bottom left panel: neurofilament light chain staining, bottom right panel: beta-amyloid precursor protein staining). (**B**) Representation of the homology model of the human V-ATPase pump and localization of the *de novo* mutations. On the left, the cytosolic V1 region (in grey ribbons) and the V0 membrane-embedded domain (the subunit a and the c-ring are in cyan and pink ribbons) are reported together with their relative functions of ATP hydrolysis and proton transport, respectively. In the centre of the figure, the G551, R740 and R804 mutations in the *ATP6V0A1* gene are represented in sticks and their position on transmembrane helix (TM) 4, 7 and 8 and with respect to the protonated residue E139 of the subunit c (V-type proton ATPase 16 kDa proteolipid subunit, NP_001685.1) is magnified on the right. S477 is localized at the exit of the luminal channel as indicated at the bottom of the figure. (**C**) Fluorescence intensity ratio of yellow (541 nm)/blue (441 nm) wavelengths of LysoSensor measured in Neuro2a cells expressing wild-type or R740Q mutant ATP6V0A1 mutant cells. (**D**) Neuro2a cells were treated with rapamycin (Rap 100 nM; 12 h) or chloroquine (CQ 50 µM; 12 h), preincubated with LysoTracker (red) and stained with Cathepsin D antibody (green) and DAPI (blue). Single channel (left), merged (centre), and inset (right) images are shown. Scale bars, 100 µm (merged) and 50 µm (inset). (**E**) Quantitative analysis of LysoTracker and Cathepsin D-positive compartments is shown. (**F**) Immunoblotting of R740Q Neuro2a mutant cells showed downregulation of the Notch signalling pathway, which relies on endosome and lysosome acidification, with activation of the cyclin-dependent kinase inhibitor p21. GAPDH is used as loading control. Abbreviations: Notch Transmembrane (NTM), Notch intracellular domain (NICD), Mastermind-like 1 (MAML1), Hairy and Enhancer of Split 1 (HES1). Displayed membranes are cropped and three independent experiments are shown. (**G**) Densitometry of the intensity of the immunoblot signals were normalised to GAPDH and expressed as fold change of R740Q mutant ATP6V0A1 samples relative to wild type. Individual data points represent independent measurements and are displayed as mean ± s.e.m. P values derived from unpaired two-tailed t-test are shown.

To provide an estimate of the frequency of *ATP6V0A1* variants, we analysed the recently published predicted damaging DNMs (n=45,221) from the largest cohort to date of exome sequence data from individuals with severe DD (n=31,058) (Kaplanis et al., 2020). We observed 11 missense mutations in *ATP6V0A1*, c.2219G>A recurring in 7 patients (6 of which are included in the present study), and concluded that *ATP6V0A1* variants are the most common DNMs among all known lysosomal disease (LD) and LSD-associated genes in this database (**Supplementary Table 1**).

Functional studies in the well characterized *Saccharomyces cerevisiae* V-ATPase (ScV-ATPase) (Roh et al., 2018) have shown that the R735 residue in subunit a, corresponding to R740 in human *ATP6V0A1*, is essential for proton transport into organelles (Kawasaki-Nishi, Nishi, & Forgac, 2001). Upon protonation of critical glutamates of subunit c, the c-ring rotates, a salt bridge with R735 is formed, and protons are transferred and pulled into the organelle lumen through a network of polar and negatively charged residues (Mazhab-Jafari et al., 2016). Homology modelling suggests that the identified *de novo* mutations overall affect organelle acidification by hindering glutamate deprotonation (p.R740Q and p.R804H), deforming the architecture of the protein region devoted to protons exchange (p.G551E), or altering the conformation of the loop that contours the exit of the luminal channel (p.S477P) (**Fig. 2B**).

To test whether V-ATPase complex function is affected by the mutations in *ATP6V0A1*, we generated Neuro2a cell lines stably expressing wild type or c.2219G>A (p.R740Q) mutant human *ATP6V0A1* and assessed acidification of the endolysosomal compartment using the LysoSensor radiometric probe, which undergoes a pH-dependent emission shift to longer wavelengths in acidic environments. Compared to wild-type, we observed a decrease in red-shifted fluorescence signal in mutant ATP6V0A1-expressing cells, indicative of impaired protonation (**Fig. 2C**). Co-localization of lysosomes with Cathepsin D, an enzyme whose trafficking to the endolysosomal compartment and maturation into an active lysosomal aspartyl protease requires an acidic pH (Braulke & Bonifacino, 2009; Ghosh, Garde, & García, 2003; Isidoro, Grässel, Baccino, & Hasilik, 1991), was nearly completely abolished by the mutant ATP6V0A1 (**Fig. 2D and E**). Furthermore, western blot analyses showed decreased proteolytic maturation of Preprocathepsin into the active 31kD form upon autophagy induction with serum starvation or rapamycin treatment in mutant cells compared to wild type (**Supplementary Fig. 3A and B**). Intriguingly, mutations resulting in the inactivation or mislocalization of Cathepsin D lead to neuronal ceroid lipofuscinosis, suggesting a convergent pathogenic mechanism with other LSDs (Siintola et al., 2006; Steinfeld et al., 2006). Autophagosome turnover, a process dependent on vacuolar acidification (Yim & Mizushima, 2020), was also impaired in cells expressing mutant ATP6V0A1, as suggested by the increased ratios of LC3-II to LC3-I and LC3-II levels alone upon autophagy induction, which persisted after removal of rapamycin (**Supplementary Fig. 4A and B**). We further investigated the impairment of endolysosomal acidification in mutant cells, by assessing Notch signalling, a critical pathway for many cellular processes (Mašek & Andersson, 2017) and dependent on V-ATPase proton pump activity for its maturation (Baron, 2012; Le Borgne, 2006; Windler & Bilder, 2010). We observed downregulation of Notch signalling in R740Q cells, with selective reduced expression of Notch target HES-1 and not of the transcriptional coactivator MALM1, and consequent de-repression of cell cycle inhibitor p21 (**Fig. 2F and G**) (Guiu et al., 2013; Kabos, Kabosova, & Neuman, 2002).

**Figure 3:**
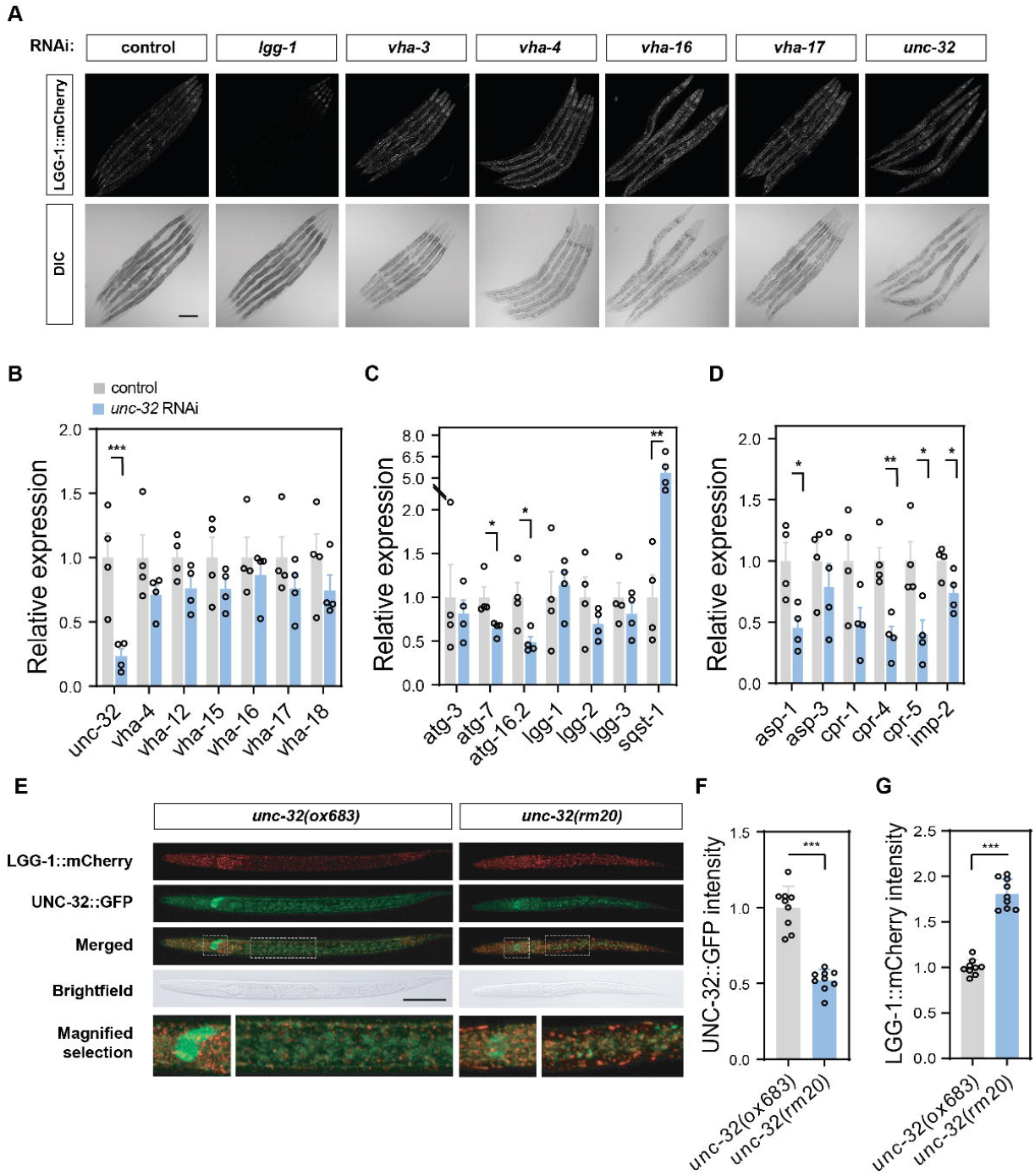
Loss of the V-ATPase a1 subunit *unc-32* causes widespread autophagy defects in *C. elegans*. (**A**) Confocal micrographs of age-synchronized adult animals expressing mCherry-tagged LGG-1 after treatment with RNAi targeting *lgg-1*, indicated V-ATPase genes or empty vector control. Scale bar, 200 µm. (**B-D**) Expression analysis of indicated genes relative to *cdc-42* in wild-type animals grown on *unc-32* RNAi normalized to vector control. (**B**) Expression level of V-ATPase subunit genes belonging to the V0 (*unc-32, vha-4, vha-16, vha-17*) and the V1 domain (*vha-12, vha-15, vha-18*) after *unc-32* RNAi treatment. (**C**) Expression level of genes belonging to the autophagic machinery after *unc-32* RNAi treatment. (**D**) Expression level of lysosomal enzymes in animals treated with *unc-32* RNAi. Individual data points represent independent measurements and are displayed as mean ± s.e.m. P values derived from unpaired two-tailed t-test are indicated with one or more asterisks (* P<0.05, ** P<0.01, *** P<0.001). (**E**) Representative confocal micrographs of *unc-32(ox683) and unc-32(rm20)* larvae expressing GFP-tagged wild-type or mutant UNC-32, respectively, as well as LGG-1::mCherry. Animals were imaged 24 hours post egg-lay, and the head and centre region are shown as magnified selection (dotted lines). Scale bar, 50 µm. (**F**) Quantification of UNC-32::GFP fluorescence intensity levels at the nerve ring in *unc-32(ox683)* and *unc-32(rm20)* animals. (**G**) Quantification of LGG-1::mCherry fluorescence intensity levels in *unc-32(ox683)* and *unc-32(rm20)* animals. Individual data points represent 9 animals from 2 independent experiments and are displayed as mean ± s.e.m. P value derived from unpaired two-tailed t-test is reported with asterisks (*** P<0.001).

We turned to the nematode *Caenorhabditis elegans* to extend our observations in an animal model of LSD (de Voer, Peters, & Taschner, 2008). Among the genes coding for V-ATPase subunit a in *C. elegans, unc-32* is most closely related to human ATP6V0A1 at protein level (Oka, Toyomura, Honjo, Wada, & Futai, 2001) (56% sequence identity and 69% similarity; **Fig. 1C**). *unc-32* is expressed throughout development and adulthood (Lynch, Briggs, & Hope, 1995; Pujol, Bonnerot, Ewbank, Kohara, & Thierry-Mieg, 2001) in multiple tissues, with highest levels in neurons (Lynch et al., 1995; Pujol et al., 2001). To evaluate the role of *unc-32* in lysosomal function, we engineered a strain to express endogenous LGG-1 (the worm ortholog of Atg8/LC3) fused to mCherry red fluorescent protein using CRISPR/Cas9. Consistent with a role of the V-ATPase complex in the autophagic pathway, RNA interference (RNAi) targeting unc-32 or other V0 subunits (*vha-3, vha-4, vha-16*, and *vha-17*) increased LGG-1::mCherry fluorescence levels relative to control (**Fig. 3A**). Knock-down of *unc-32*, without altering the levels of other subunits of the V-ATPase complex (**Fig. 3B**), reduced the expression of components of the autophagic machinery (*atg-7, atg-16*.*2*) (**Fig. 3C**) and lysosomal hydrolytic enzymes (*asp-1, cpr-4, cpr-5, imp-2*) (**Fig. 3D**), and increased expression of the stress-responsive autophagy receptor *sqst-1* (**Fig. 3B**). Interestingly, *lgg-1* expression was unchanged by *unc-32* RNAi, indicating that the observed increase of LGG-1::mCherry levels (**Fig. 3A**) is caused by reduced clearance of this protein by the lysosome. We also performed Cas9-mediated editing of *unc-32* in a strain that has *gfp* inserted at the endogenous locus to introduce a point mutation corresponding to R740Q in human ATP6V0A1 (**Supplementary Fig. 5A, B and C**). Animals homozygous for the mutant *unc-32(rm20)* allele show developmental arrest at early larval stages (**Fig. 3E**), reduced levels of UNC-32::GFP at the nerve ring (**Fig. 3F**), as well as a widespread accumulation of enlarged LGG-1::mCherry puncta (**Fig. 3G**), further demonstrating the essential role of V-ATPase activity in organismal health.

## Discussion

The eukaryotic V-ATPase is a multi-subunit rotary pump, consisting of a soluble catalytic V1 and a membrane-bound V0 components, which mediates the acidification of cellular and intracellular compartments by driving proton translocation across the membrane (Collins & Forgac, 2020). Over the last years, a number of non-canonical functions of the V-ATPase have been unravelled, which are distinct but functionally related with its primary proton pump role, including membrane fusion, nutrient sensing, and scaffold for protein-protein interactions (Hiesinger et al., 2005; Hurtado-Lorenzo et al., 2006; Zhang et al., 2014). Isoforms of many of the V1 and V0 subunit proteins are encoded by multiple genes or alternatively spliced transcript variants, with tissue specific expression and mechanisms of regulation (Collins & Forgac, 2020). The *ATP6V0A1* gene encodes the a1 isoform, which of the four a-subunits is the most neuronal enriched, mainly localising to the nerve terminals (Morel, Dedieu, & Philippe, 2003).

Recently, Aoto et al. identified two subjects with intellectual disability and epilepsy carrying a *de novo* missense R741Q substitution in *ATP6V0A1* and two with biallelic variants, comprising one complete loss of function and the A512P or N534D missense variant (Aoto et al., 2021), respectively corresponding to R740Q, A505P and N527D in the updated accession number NM_001130021.3 used in this manuscript. Here we report 17 subjects from 14 unrelated families carrying the R740Q, other newly identified *de novo* missense or inherited biallelic variants in this gene, representing the largest cohort of *ATP6V0A1*-associated disease mutations to date. With 58 DNMs (50 missense, 3 indel, 2 splice site, 2 initiator codon, 1 frameshift) in genes of the V-ATPase complex identified in exome sequence data from over 30,000 individuals with severe DD (Kaplanis et al., 2020), and significant association with severe DD for 2 genes of this group (*ATP6V0A1* and *ATP6V1A*) out of 281, we propose that a V-ATPase-related disease should be considered in individuals presenting with mild-to-profound developmental delays and epilepsy. Using *in vitro*, computational and *C. elegans* modelling, we next proceeded to study the mechanism of pathogenicity of the R740Q variant, which is the most prevalent of all the *de novo* cases reported so far and the one resulting in the most severe phenotype. Our experimental work indicates that the R740Q variant causes altered organelle acidification and failure of lysosomal hydrolysis directly by impairment of the canonical proton-pumping V-ATPase function. The compromised acidification of the endolysosomal compartments leads to impaired γ-secretase-mediated processing and release of the Notch intracellular domain (NICD) and reduced expression of Notch target genes. These findings corroborate recent reports indicating that in *Drosophila* the V-ATPase-mediated acidification of the endolysosomal compartment is required for the activation of Notch in endosomes (Sorensen & Conner, 2010) and expression of a dominant negative subunit of V-ATPase in neural precursors reduced Notch signaling and depleted neural stem cells leading to neuronal differentiation (Lange et al., 2011). The severe impairment of the autophagic machinery upon knock down of the *ATP6V0A1* ortholog in *C. elegans*, and the reduced protein clearance by lysosomes upon introduction of the R740Q variant in homozygote state reported here, together with the recent description of severe impairment of the lysosomal dysfunction and decreased Cathepsin D activity in homozygous mutant mice harbouring the human R741Q and A512P variants (Aoto et al., 2021), recapitulate the results observed in neuronal cell lines upon overexpression of the R740Q variant, supporting an antimorphic mode of action of the R740Q and the other *de novo* variants.

Five individuals from 2 unrelated pedigrees in our cohort carry the biallelic E149Kfs*18 frameshift and R495W substitution and manifest an early onset, progressive myoclonus epilepsy and ataxia, therefore expanding the neurological phenotypes associated with *ATP6V0A1* variants beyond the more severe developmental and epileptic encephalopathy observed in the *de novo* cases. Of note, the parents carrying only one of the variants do not manifest any obvious neurological phenotype, suggesting a mechanism of haploinsufficiency for the inherited variants. The R495W substitution results in loss of a positively charged hydrophobic residue in close proximity of S477 and the luminal channel exit. We speculate that this variant has a less damaging impact on the proton pumping activity, leading to neurological manifestations only when in compound heterozygosity with a null allele. Further work is needed to clarify the precise molecular mechanisms through which the described biallelic and *de novo* mutations result in a PME and DEE phenotype, respectively. Considering the increasing identification of disease-causing variants in V-ATPase genes and the critical relevance of vacuolar pump-mediated regulation of intracellular pH in cellular homeostasis in health and disease (Vasanthakumar & Rubinstein, 2020), this multi-protein complex is rapidly taking centre stage as a molecular hub for unravelling the disease mechanisms of LDs and other human diseases, from cancer to neurodegeneration.

## Materials and methods

### Patients

Ethical approval was obtained from the ethics committee of the following institutes: University of Federico II (Italy), University of L’Aquila (Italy), G. Gaslini Institute (Italy), University of Exeter (UK), Nottingham University Hospital (UK), Salpêtrière Hospital (France), University of Toronto (Canada), University of Ottawa (Canada), St. Luke’s Children’s Hospital (USA), Floating Hospital for Children at Tufts Medical Center (USA), Gillette Children’s Specialty Healthcare Children’s Hospital (USA), Children’s Hospital of Philadelphia (USA), Peking University First Hospital, Beijing (China). Written consent for the study was obtained from patients or legal representatives according to the Declaration of Helsinki. When individuals were not contacted directly, de-identified phenotypic and genomic data were used. Patient or guardian consent was given for the publication of the patient’s data and clinical information. Patient II.1 was enrolled in a research study aimed at finding the genetic cause of progressive myoclonus epilepsy; Patient III.1, VII.1-X.1, and XIV.1 were referred to GeneDx for clinical whole-exome sequencing for diagnosis of suspected Mendelian disorders as previously described (Retterer et al., 2016); Patient IV.1 was recruited in the Italian Undiagnosed Rare Diseases Network (IURDN; PGR00229, 2016–19); Patient V.1 and VI.1 were part of the Deciphering Developmental Disorders Study (DDD; 10/H0305/83). Patient XI.1 was recruited as part of the Care4Rare Canada research study.

### Sequencing and genotyping

Blood samples were collected and DNA was extracted using standard methods from peripheral blood lymphocytes from the indicated individuals in Figure 1A.

Fam. I: Exome sequencing was performed as previously described (Rinaldi et al., 2015). Libraries were prepared using the SeqCap EZ Human Exome Library version 2.0 (Roche Nimblegen Inc) and a 100–base pair paired-end run was performed on the HiSeq 2000, with each sample on a single lane of a TruSeq version 2 flow cell (Illumina). Sequence alignment, quality control, and variant calling were performed with BWA, SAMTools, the Genomic Analysis Toolkit, and Picard (http://picard.sourceforge.net/index.shtml). Data analysis was based on the autosomal recessive mode of inheritance. Variant calling and quality score recalibration was performed using GATK (http://www.broadinstitute.org/gatk/). Variants remaining after exome data analysis containing missing data were Sanger sequenced using the BigDye Terminator version 3.1 chemistry (Applied Biosystems), run on an ABI 3730xl analyzer, and analyzed using Sequencher software version 4.2 (Gene Codes Corporation). Fam. II-XIV: Details on sequencing, alignment, variant calling (inherited and *de novo*) and variant annotation have been described previously (Kaplanis et al., 2020; Kinay et al., 2018; Mazzola et al., 2020; McRae et al., 2017; Muona et al., 2015; Niestroj et al., 2020; Salvatore et al., 2020). Presence or absence of the disease-causing variants was confirmed on DNA of the proband and additional members from each family by Sanger sequencing.

### Homology modelling and structure analysis of the human V-ATPase

Homology models of the subunit a (NM_001130021.3, NP_001123493.1) and c (NM_001694.4, NP_001685.1) belonging to the V0 domain of the human V-ATPase has been built by Robetta server (Kim, Chivian, & Baker, 2004), using as templates the 3D structures of the homologous subunits in *Bos Taurus* (Wang et al., 2020) (PDB ID: 6XBW; 97% and 98% identity between the homologous subunits a and c). The 3D models of the G551E and S477P mutants has been built with Robetta (Kim et al., 2004). Structural analysis has been carried out with Coot and Pymol (The PyMOL Molecular Graphics System, Version 1.2r3pre, Schrödinger, LLC).

### cDNA cloning and generation of stable cell lines

Human *ATP6V0A1* cDNA was kindly provided by Michael Forgac. Point mutations corresponding to the gene variants identified in the present study were introduced using a QuikChange II site-directed mutagenesis kit (Agilent Technologies) according to the manufacturer’s instructions, and subsequently the constructs were cloned into the PiggyBac (PB) Transposon vector clone (Stratech, PB511B-1). For stable transfection, PB Transposon vector and PB Transposase vector (Stratech, PB210PA-1) were transfected at a ratio of 9:1 into Neuro2a cells plated on 24-well plates (∼50,000 cells per well). One day after transfection, the cells were trypsinized and transferred as serial dilution to fresh tissue culture plates. Drug selection using 10 µg/mL puromycin started on day 3 and was continued for 2 to 3 weeks until the visible colonies appeared. In order to induce the transgene expression, 1 µg/mL doxycycline was added to the media starting 1 day before transfection.

### Immunofluorescence

After fixation with 4% paraformaldehyde, slides were placed in blocking solution (10% normal goat serum and 0.3% Triton X-100 in phosphate-buffered saline [PBS]) for 45 minutes at room temperature. Primary antibody staining was done at 4°C overnight in PBS with 5% normal goat serum and 0.1% Triton X-100 (The Dow Chemical Company), using LysoTracker fluorescent dye (L12492, ThermoFischer Scientific) and Cathepsin D antibody (2284, Cell signaling). The slides were then washed 3 times with PBS (0.1% Triton X-100 in PBS), incubated with secondary antibody (Invitrogen, 1:500) for 1 hour at room temperature in the dark, and then washed 3 times before drying and adding Vectashield/4′,6-diamidino-2-phenylinodole stain (Vector Laboratories). Coverslips were mounted with Permount (Fisher Scientific). The cells were imaged and acquired using the PANNORAMIC 250 fluorescent microscope scanner (3DHISTECH). An average of 100 cells from 3 independent experiments per each condition were analysed by a blinded investigator.

### Endolysosome pH measurement

Quantification of lysosomal pH was determined using Dextran conjugated Lysosensor Yellow/Blue DND-160 (L7545, Invitrogen). Wild-Type and R740 mutant Neruo2a cells were grown in serum starved conditions (DMEM with antibiotics) with 1 µg/mL doxycycline to ∼80% confluence. Cells were then trypsinized, harvested and aliquoted at 100 µl into a black 96-well microplate with 5µM Lysosensor probe for 15 min at 37°C with 5% CO2 (50,000 cells). The cells were then washed 3X in DMEM and the samples were read in a CLARIOstar Plus spectrophotometer (BMG Labtech) with dual-excitation and dual-emission spectral peaks at 324/441 and 381/541 nm. The ratio of emission was then calculated for each sample as indicated.

### Western blotting and densitometry analysis

Whole cell lysates were collected as previously described (Forouhan, Mori, & Boot-Handford, 2018). Protein concentration of whole cell lysates were determined using the Pierce bicinchoninic acid protein assay (23227, Thermo Scientific) with a bovine serum albumin standard curve according to the manufacturer’s protocol. Twenty micrograms of denatured protein were loaded into the precast 4–12% Bis-Tris gels (NP0322BOX, Life Technologies) or 16% Tricine gel (EC6695A, Thermo Scientifc) for LC3 immunoblot. The gel was electroblotted onto 0.45 μm nitrocellulose membrane or 0.2 μm PVDF membrane for LC3 western blotting, which was then blocked for 1 h at room temperature with 5% skim milk in PBS containing 0.1% (v/v) Tween-20 and 2% (v/v) serum. The membranes were incubated overnight at 4°C in 1/1000 dilution of following primary antibodies; Notch isoforms using Notch isoform sampler kit (3640, Cell Signalling), Cathepsin D antibody (2284, Cell signaling), LC3 A/B (12741, Cell Signalling), and GAPDH (sc-47724), and HRP-linked secondary antibodies. Blots were visualized with chemiluminescence reagent (Life Technologies). Densitometric quantification of bands was performed with the ImageJ software and standardized relative to a loading control against a control protein sample on each blot. Ratios were normalized for each individual experiment, with the wild type sample set as 1.

### *C. elegans* strains and maintenance

*C. elegans* were maintained on solid nematode growth medium seeded with *E. coli* OP50 at 20 °C using standard methods (Brenner, 1974). Strains used in this study were Bristol N2 and EG9591 [*unc-32(ox683[unc-32::gfp +loxP])* III] (Schwartz & Jorgensen, 2016).

The insertion of *mcherry* at the 5’ end of endogenous *lgg-1*, and the introduction of a missense mutation corresponding to the substitution of the essential arginine by a glutamine in UNC-32::GFP at position 804 (numbered according to the isoform UNC-32A) into the EG9591 strain, were performed using CRISPR/Cas9 genome editing and resulted in strains AM1219 *lgg-1(rm17[lgg-1::mcherry])* II and AM1232 *unc-32(rm20[unc-32(R804Q)::gfp +loxP])* III*/hT2[bli-4(e937) let-?(q782) qIs48]* (I;III).

### CRISPR/Cas9-mediated genome editing

Genome editing was performed as previously described (Paix, Folkmann, & Seydoux, 2017). Briefly, Cas9 ribonucleoprotein complexes were assembled in vitro from purified Cas9 (New England Biolabs), tracrRNA and gene specific crRNA (Integrated DNA Technologies), and injected together with a linear DNA repair template in the gonad of adult hermaphrodites. CRISPR editing was performed using *dpy-10* to generate *cn64* rollers as phenotypic marker. Successful edits were identified by PCR screening and verified by Sanger sequencing.

For inserting *mcherry* at the endogenous *lgg-1* locus, we targeted the 5’ end of the first exon with the guide sequence 5’-CCTTCGAATCAAAATGAAGT-3’. The repair template was synthesized by PCR to generate a double-stranded DNA fragment consisting of mCherry amplified from pAP582 (Addgene) and homology overhangs for insertion at the ATG start site using primers 5’-TAACCTTCTCTTCACACTAACCTTCGAATCAAAATGGTCTCAAAGGGTGAAGAAGATAAC -3’ and 5’-CTTCTCAAAGTTGTTCTCCTCCTTGTAAGCCCACTTCTTATACAATTCATCCATGCC-3’.

The *lgg-1(rm17)* allele was isolated in a dpy-10(+) background and the resulting strain was back-crossed three times to N2.

For generating the R804Q substitution in UNC-32::GFP, we edited the *unc-32(ox683)* allele using the guide sequence 5’-CTGCTTCATACCTTCGTCTT-3’ and a single-stranded oligonucleotide containing the CGT>CAG change, a silent T>G mutation resulting in the PvuII restriction site for genotyping, and homology arms flanking the edit as repair template 5’-TCTTGGATGTGTGTCACATACTGCTTCATACCTTCagCTgTGGGCTCTTTCATTGGCTCATGCTCGTAAGTAAAG-3’. The resulting *unc-32(rm20)* allele was confirmed by sequencing and kept on the *hT2* balancer for strain maintenance.

### RNA interference

RNAi-mediated knock-down of V-ATPase genes or *lgg-1* was performed by feeding animals with *E. coli* strain HT115(DE3) containing the appropriate RNAi vectors obtained from the Ahringer library and using L4440 as the empty vector control (Kamath & Ahringer, 2003). Bacterial cultures were grown overnight in LB with 100 ug/mL ampicillin and induced with 5 mM IPTG for 3 hours at 37 °C before plating. Animals were age-synchronized by a 2-hour egg-lay on RNAi plates and collected for analysis 72-96 hours later. For RNAi clones that cause developmental defects (*vha-4, vha-16*, and *vha-17*), the animals were initially grown on L4440 and transferred to RNAi plates at L4 stage.

### Fluorescence imaging

Animals were immobilized in 50 mM sodium azide on 3% agarose pads and imaged using a Zeiss LSM800 confocal microscope with 10x or 20x objective and Zen imaging software. For quantification of LGG-1::mCherry fluorescence levels, maximum intensity projections were generated from Z stack images and fluorescence intensity was quantified by tracing either the nerve ring for UNC-32::GFP or whole animals for LGG-1::mCherry using ImageJ software (NIH).

### Gene expression analysis in *C. elegans*

Age-synchronized populations of at least 100 gravid adults were collected and snap frozen in liquid nitrogen. RNA was extracted using TRIzol (Invitrogen) and purified with QIAGEN RNeasy MinElute columns as per manufacturers’ instructions (Sala, Bott, Brielmann, & Morimoto, 2020). mRNA was reverse transcribed using the iScript cDNA Synthesis Kit (BioRad) and real-time quantitative PCR was performed using iQ SYBR Green Supermix (BioRad) in a BioRad CFX384 Real-Time PCR system. Relative expression was determined from cycle threshold values using the standard curve method and the expression of genes of interest was normalized to *cdc-42*. The primers used are listed in Supplementary Table 2.

### Statistical analysis

Statistical analysis was performed with Graph Pad Prism version 8 for Windows (GraphPad Software, San Diego California USA, www.graphpad.com), using the tests indicated in the figure legends. A standard confidence interval of 95% was applied in all analyses. Displayed in the figure are the mean values of all technical replicates for each of the independent experiments (displayed as single data points). Error bars represent the standard error of the mean. P values <0.05 were considered statistically significant.

## Data Availability

All data needed to evaluate the conclusions in the paper are present in the paper and/or the Supplementary Materials.

## Data availability

The raw data that support the findings of this study are available from the corresponding authors, upon request.

## Acknowledgements

We are grateful to the participants and the core research staff who made this study possible. We thank Karen L Oliver (University of Melbourne) for the project and case management in the progressive myoclonus epilepsy exome study. We thank Dr. Bryan J. Traynor (National Institute on Aging, NIH) for the help with the exome sequencing of Fam. 1. We thank Michael Forgac for ATP6V0A1 plasmid constructs, the Caenorhabditis Genetics Center and Erik Jorgensen for *C. elegans* strains, Jian Li and John Rubinstein for valuable discussions, and Renee Brielmann for technical assistance in this project. We also thank the High-Throughput Analysis Lab, the Keck Biophysics Facility, and the laboratory of Robert Lamb at Northwestern University for instrument use.

## Funding

This research was supported in part by the Intramural Research Program of the National Institutes of Health, National Institute of Neurological Disorders and Stroke, grant Z01-AG000949-02 from the National Institute on Aging (Dr Johnson). The Italian Network of Rare Diseases is funded by the Italian Ministry of Foreign Affairs and International Cooperation and Farmindustria (PGR00229, 2016–19). The Care4Rare Canada Consortium is funded by Genome Canada and the Ontario Genomics Institute (OGI-147), the Canadian Institutes of Health Research, Ontario Research Fund, Genome Alberta, Genome British Columbia, Genome Quebec, and Children’s Hospital of Eastern Ontario Foundation. L.C.B. was supported by postdoctoral fellowships from the National Ataxia Foundation and the American Federation for Aging Research, as well as a research grant from the Kennedy’s Disease Association. M.F. and C.R. are supported by the Wellcome Trust under fellowship award 205162/Z/16/Z. W.F.L. is supported by a research grant from the Kennedy’s Disease Association and a John Fell Fund from the Medical Science Division of the University of Oxford. R.I.M. received support from the National Institutes of Health (National Institute on Aging R56AG059579, R37AG026647, RF1AG057296, and P01AG054407) and the Daniel F. and Ada L. Rice Foundation. P.S. developed this work within the framework of the DINOGMI Department of Excellence of MIUR 2018-2022 (legge 232 del 2016).

## Competing interests

The authors report no competing interests.

## Supplementary material

Supplementary material is available online.

## Appendix 1

Appendix contains the list of consortium collaborators.

## Notes

### Competing Interest Statement

The authors have declared no competing interest.

### Funding Statement

All funding support in the present work is disclosed in the manuscript.

### Author Declarations

Ethical approval was obtained from the ethics committee of the following institutes: University of Federico II (Italy), University of L Aquila (Italy), G. Gaslini Institute (Italy), University of Exeter (UK), Nottingham University Hospital (UK), Salpetriere Hospital (France), University of Toronto (Canada), University of Ottawa (Canada), St. Luke s Children's Hospital (USA), Floating Hospital for Children at Tufts Medical Center (USA), Gillette Children s Specialty Healthcare Children's Hospital (USA), Children s Hospital of Philadelphia (USA), Peking University First Hospital, Beijing (China). Written consent for the study was obtained from patients or legal representatives according to the Declaration of Helsinki. When individuals were not contacted directly, de-identified phenotypic and genomic data were used. Patient or guardian consent was given for the publication of the patient s data and clinical information.

