## Supplementary Material for "Biallelic and *de novo* variants in *ATP6V0A1* cause progressive myoclonus epilepsy and developmental and epileptic encephalopathy"

**SUPPLEMENTARY FIGURES**

**Supplementary Figure 1: Identification of the disease-causing mutation in Family 1.** The filtering paradigm used to detect novel sequence variants that co-segregated with disease is shown. SNV: single nucleotide variant.

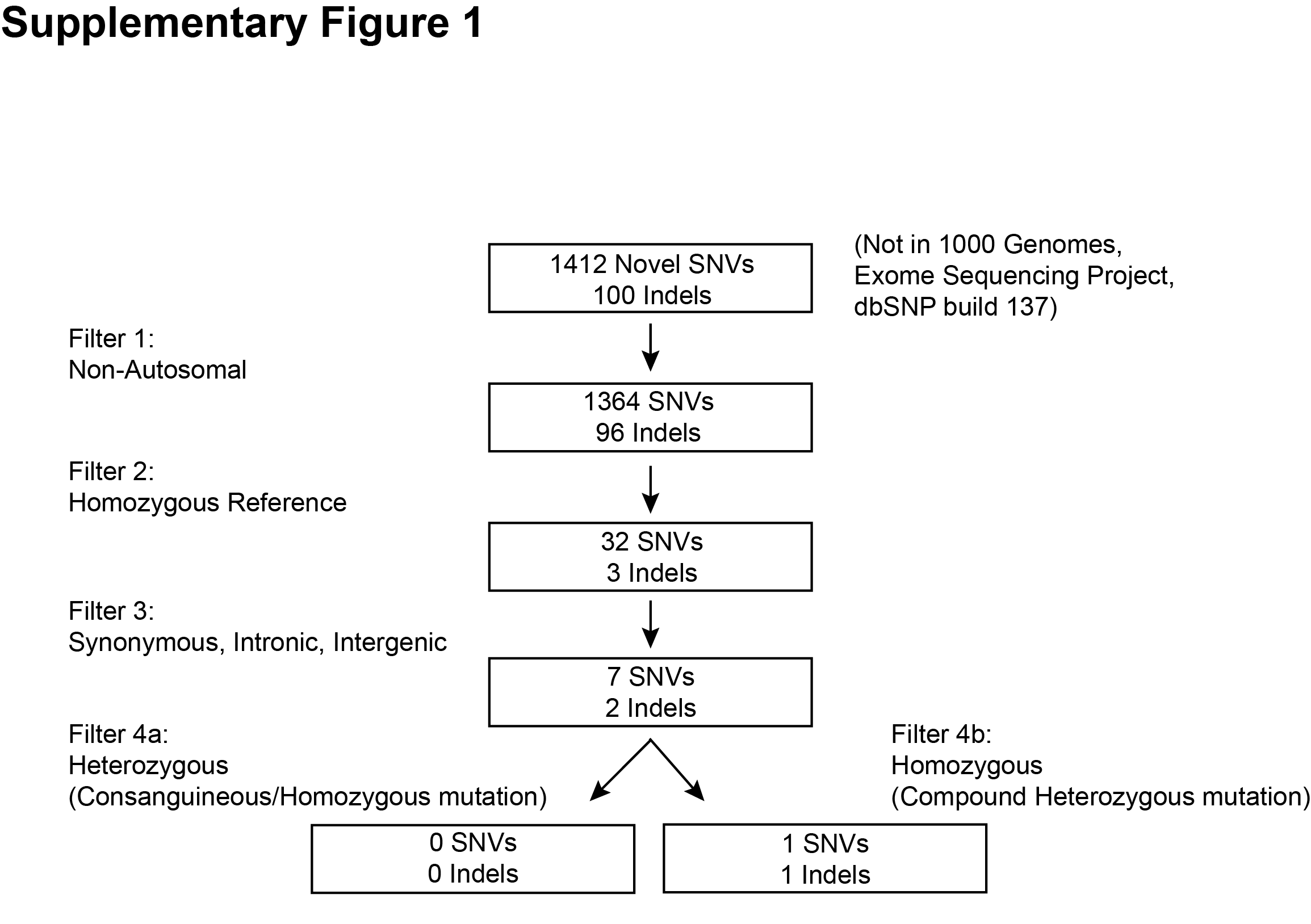

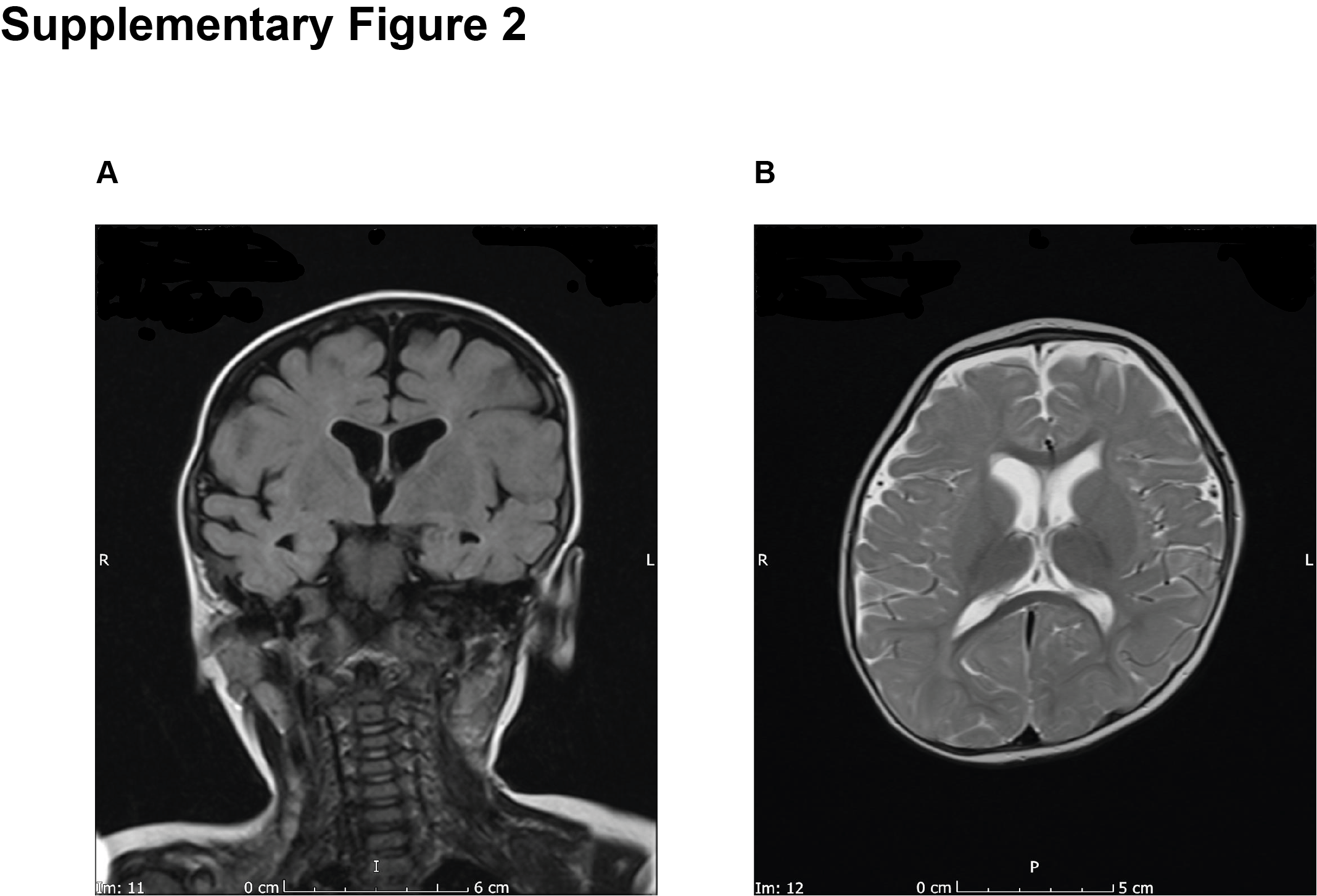

**Supplementary Figure 2: Representative brain MRI images of P:V.1 carrying the c.2352G>A mutation in the *ATP6V0A1* gene.** (**A**) Coronal FLAIR and (**B**) and axial T2-weighted brain MR images showed deformed skull and brain parenchyma, in absence of focal brain lesion. Occipito-frontal head circumference was below the 1^st^ centile for age and sex.

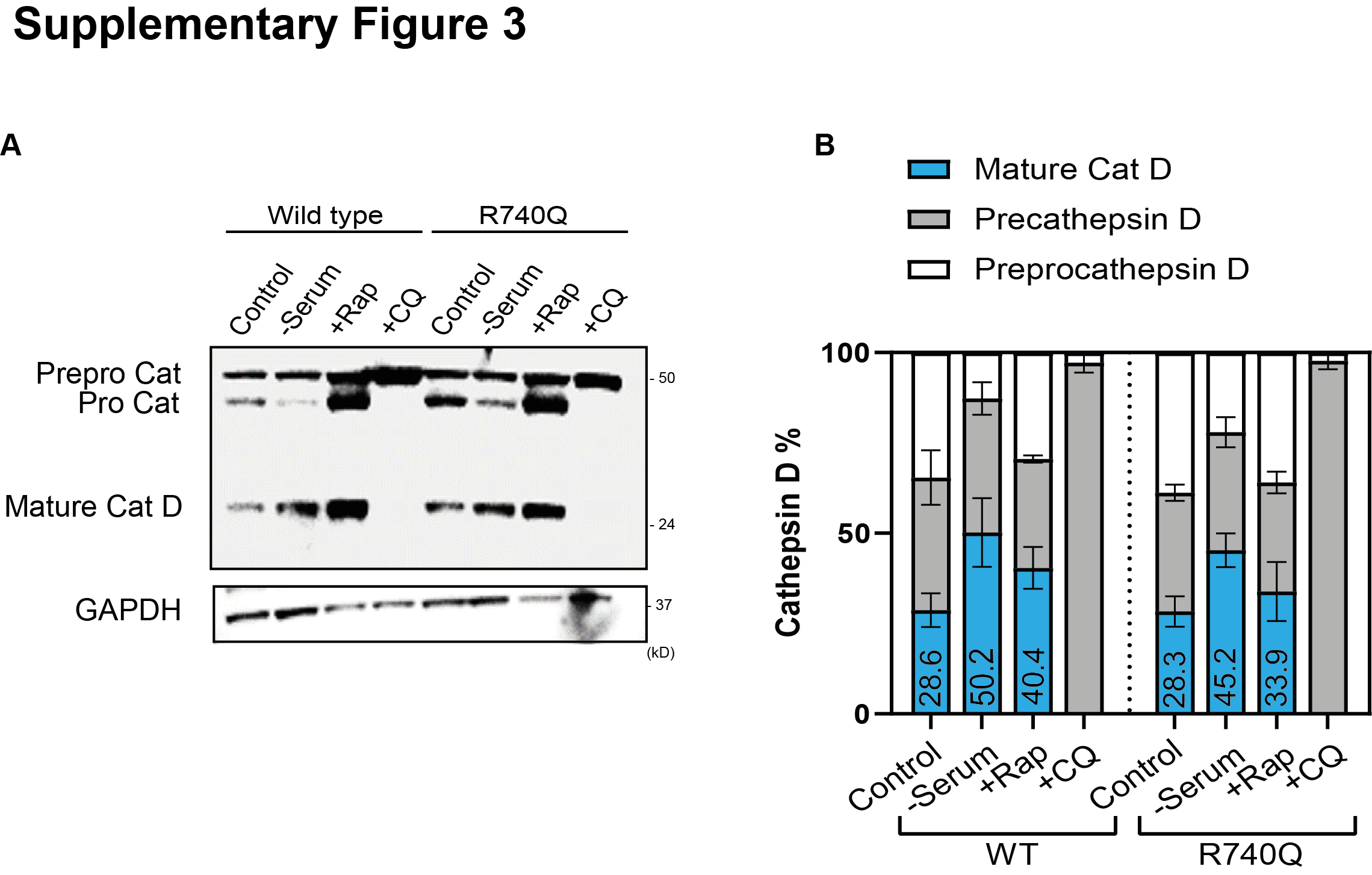

**Supplementary Figure 3: Cathepsin maturation is impaired in ATP6V0A1 mutant cells.** (**A**) Immunoblotting shows Cathepsin D (Cat D) processing in R740Q mutant compared to wild type Neuro2a cells, under conditions of no treatment (control), autophagy induction with serum starvation (- serum) and rapamycin (Rap 100 nM; 12 h), and alkalinisation of endolysosomal vacuoles using chloroquine (CQ 50 µM; 12 h). GAPDH is used as loading control. Displayed membranes are cropped and one representative out of three independent experiments is shown. (**B**) Densitometry of the intensity of the immunoblot signals were normalised to GAPDH and expressed as percentage of the total Cathepsin D signal. Average percentages of mature Cat D are shown. Values are displayed as mean ± s.e.m.

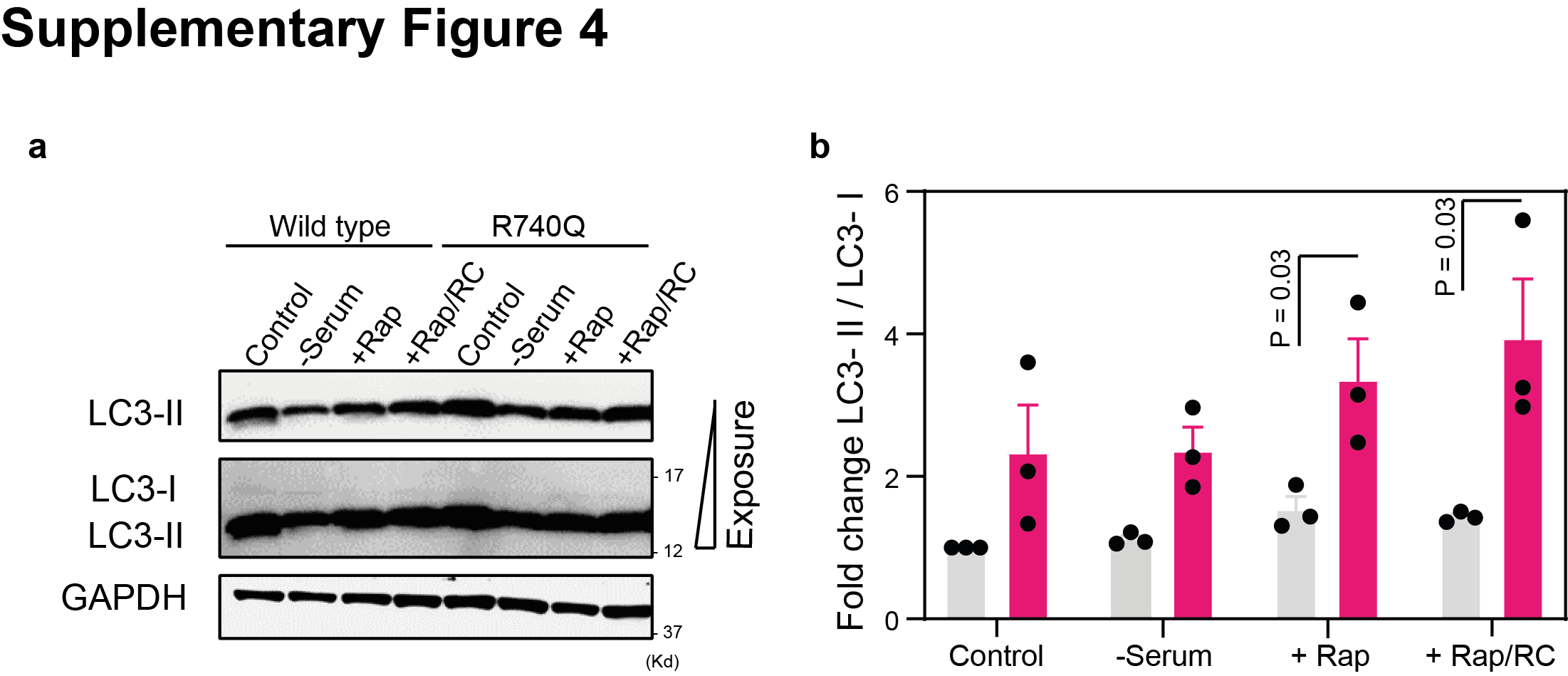

**Supplementary Figure 4:** **Impaired Clearance of LC3-II from Autolysosomes in ATP6V0A1 mutant cells.** (**A**) Immunoblotting of wild type and R740Q Neuro2a mutant cells in cells under conditions of no treatment (control), serum starvation (- serum), rapamycin (Rap 100 nM; 12 h), or rapamycin treatment followed by rapamycin removal (RC) (Rap 100 nM; 12 h) is showed. GAPDH is used as loading control. LC3: Microtubule‑associated protein light chain 3. Displayed membranes are cropped and one representative out of three independent experiments is shown. (**B**) Densitometry of the intensity of the immunoblot signals were normalised to GAPDH and expressed as fold change of R740Q mutant ATP6V0A1 samples relative to wild type. Individual data points represent independent measurements and are displayed as mean ± s.e.m. P values derived from paired two-tailed t-test are shown.

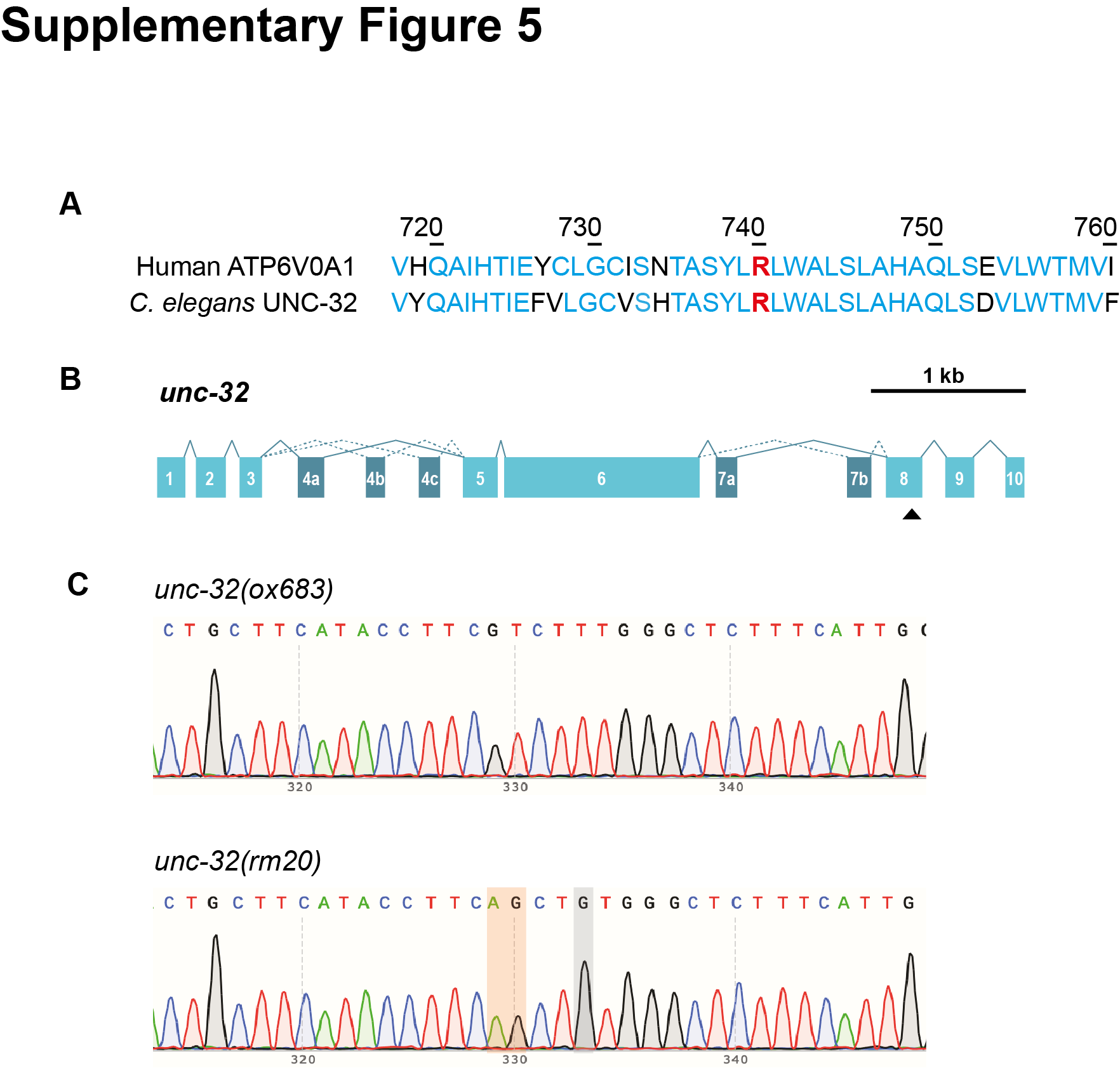

**Supplementary Figure 5:** **Unc-32 is the *C. elegans* ortholog of ATP6V0A1.** (**A**) Alignment of the amino acid sequence flanking the essential arginine that is required for proton translocation in human ATP6V0A1 and *C. elegans* UNC-32. Common residues are shown in blue. R740 in human ATP6V0A1 (corresponding to R804 in *C. elegans*) is highlighted in red. Numbers indicate amino acid position of human ATP6V0A1. (**B**) Schematic structure of the *unc-32* gene is displayed. Numbered boxes indicate exons, which are connected by a continuous line. Alternative isoforms result from inclusion or exclusion of alternative exons (dark blue), connected by dotted lines. Black arrowhead indicates the position encoding R804. (**C**) Representative electropherograms showing base substitutions (highlighted in the red box) and flanking regions in the *C. elegans unc-32(ox683)* gene (top), resulting in the *unc-32(rm20)* allele (bottom) and arginine to a glutamine change at position 804 in UNC-32 protein. The grey box is a silent mutation to create a restriction site (PvuII) that was used to identify successful edits.

**SUPPLEMENTARY TABLE**

**Supplementary Table 1.** Mutation frequency of LSD-associated genes in the DD database

| Gene ID | Mutation count | Mutation frequency in DDs |
| --- | --- | --- |
| *ATP6V0A1* | 11 | 0.00035418 |
| *MYO5A* | 6 | 0.00019319 |
| *LYST* | 6 | 0.00019319 |
| *MAN2B1* | 6 | 0.00019319 |
| *CLCN7* | 4 | 0.00012879 |
| *HPS5* | 4 | 0.00012879 |
| *HEXA* | 4 | 0.00012879 |
| *GALC* | 4 | 0.00012879 |
| *GRN* | 3 | 9.6593E-05 |
| *ATP13A2* | 3 | 9.6593E-05 |
| *SCARB2* | 3 | 9.6593E-05 |
| *NPC1* | 3 | 9.6593E-05 |
| *IDUA* | 3 | 9.6593E-05 |
| *SGSH* | 3 | 9.6593E-05 |
| *ARSB* | 3 | 9.6593E-05 |
| *GBA2* | 2 | 6.4396E-05 |
| *ARSA* | 2 | 6.4396E-05 |
| *IDS* | 2 | 6.4396E-05 |
| *LAMP2* | 2 | 6.4396E-05 |
| *PPT1* | 2 | 6.4396E-05 |
| *CLN8* | 2 | 6.4396E-05 |
| *CTSF* | 2 | 6.4396E-05 |
| *HPS3* | 2 | 6.4396E-05 |
| *HPS4* | 2 | 6.4396E-05 |
| *HPS6* | 2 | 6.4396E-05 |
| *RAB27A* | 2 | 6.4396E-05 |
| *GNPTAB* | 2 | 6.4396E-05 |
| *CTNS* | 2 | 6.4396E-05 |
| *SLC17A5* | 2 | 6.4396E-05 |
| *PPT1* | 2 | 6.4396E-05 |
| *MANBA* | 2 | 6.4396E-05 |
| *AGA* | 2 | 6.4396E-05 |
| *NAGA* | 2 | 6.4396E-05 |
| *GALNS* | 2 | 6.4396E-05 |
| *CLN3* | 1 | 3.2198E-05 |
| *MFSD8* | 1 | 3.2198E-05 |
| *KCTD7* | 1 | 3.2198E-05 |
| *HPS1* | 1 | 3.2198E-05 |
| *GNPTG* | 1 | 3.2198E-05 |
| *MCOLN1* | 1 | 3.2198E-05 |
| *CTSA* | 1 | 3.2198E-05 |
| *LIPA* | 1 | 3.2198E-05 |
| *HGSNAT* | 1 | 3.2198E-05 |
| *GNS* | 1 | 3.2198E-05 |
| *GLB1* | 1 | 3.2198E-05 |
| *HYAL1* | 1 | 3.2198E-05 |
| *GLB1* | 1 | 3.2198E-05 |
| *HEXB* | 1 | 3.2198E-05 |

| **Target** | **Forward (5’-3’)** | **Reverse (5’-3’)** |
| --- | --- | --- |
| *asp-1* | TGGTCCACCGAGCTTTACAC | TTGACTGGGAATTGGGCTCC |
| *asp-3* | ATCCACCTGCCTCTCTGGAT | AGAAGCGTCCGATGAAGACG |
| *atg-3* | TCCGCCGATCATTCCAACAA | AACGGTGGGCATTGTGAGAT |
| *atg-5* | CGACGACGGTGAAAAAGTGC | TCCGATAGGTTCGCTTGATCG |
| *atg-7* | GGCTTCCAACAGTTTTGGCT | CATGCCGAATGATGACGTAGG |
| *atg-16.2* | TTTGCAAGAAGAACGCGCAC | GATGCTCGCTCGTCACCTAA |
| *cdc-42* | GGTTGCTCCAGCTTCATTC | AACAAGAATGGGGTCTTTGA |
| *cpr-1* | CGGAGGACACGCCATTAAGA | CCATGAGTTGGCAACAAGCC |
| *cpr-4* | CGGAGGACATGCCATCAGAA | CCCCAGTTGACGTTCCATGA |
| *cpr-5* | AACCCCATACTGGCTTGTCG | AGTGCTCGATTCCACACTCG |
| *gfp* | CTGTCCACACAATCTGCCCT | TGCCATGTGTAATCCCAGCA |
| *imp-2* | TGCTTGGGCTTGGAGACATT | ACTCGGCAGTCGTTTGAACA |
| *lgg-1* | CGCATCCAACTTCGTCCAGA | TGTCCCATTGTGGTCATGGTT |
| *lgg-2* | AGCATTCTTCCTGCTCGTCAA | AAGCCATCTGGATCACGCTC |
| *lgg-3* | ATATCGCAGATGCGCCAGTT | AAGGAAGCAACAGTGTCCGT |
| *sqst-1* | TCGAGAGCCTCAGTCCATCA | TTCCTGGGCTTCAACAGCAA |
| *unc-32* | TGTTCGGTGATATGGGGCAC | GCCGCTTCGAGTTGTTTCTC |
| *vha-4* | TGTCTGTGGTCTTGCTGTCG | CGAAAAGAGCTGGATTGGCG |
| *vha-16* | ACTGGACTCTCTCGTGCTGA | CCAAAGTCTTCTCTCCTGGTCC |
| *vha-17* | CCACTGATCGGCCCACAAAT | TTGGTGCGTCTCCCCATTTT |
| *vha-12* | AATTCCTCGCCTACCAGTGC | AAGAGCTTCGGCGTATGAGG |
| *vha-15* | TCTTTCCGAGTCCACGAAGG | CCCTCTTGACTTCACGCTCA |
| *vha-18* | ATCACTCCATTGGTCGCCTG | AGCTTCAGAAGTTCCTGTCGG |

**Supplementary Table 2.** List of qPCR primers for *C. elegans*
