## Appendix 1 for "Biallelic and *de novo* variants in *ATP6V0A1* cause progressive myoclonus epilepsy and developmental and epileptic encephalopathy"

**Italian Undiagnosed Diseases Network:**

Domenica Taruscio^1^, Marco Salvatore^1^, Agata Polizzi^1^, Federica Censi^1^, Giovanna Floridia^1^, Giuseppe Novelli^2^, Erica Daina^3^, Alessandra Ferlini^4^, Marcella Neri^4^, Dario Roccatello^5^, Simone Baldovino^6^, Elisa Menegatti^6^

^1^Undiagnosed Rare Diseases Interdepartmental Unit, National Centre for Rare Diseases, Istituto Superiore di Sanità, 00161 Rome, Italy.

^2^Department of Biomedicine & Prevention, Genetics Section, University of Rome Tor Vergata, Rome 00133, Italy; IRCCS Neuromed, Pozzilli, Isernia 86077, Italy; Department of Pharmacology, School of Medicine, University of Nevada, Reno, NV 89557, USA.

^3^IRCCS Mario Negri Pharmacological Research Institute, Bergamo 24020, Italy.

^4^Department of Experimental and Diagnostic Medicine, University of Ferrara, Ferrara 44121, Italy.

^5^Nephrology and Dialysis Universitary Unit, and Center of Research of Immunopathology and Rare Diseases (CMID) San Giovanni Bosco Hospital, and Department of Clinical and Biological Sciences, University of Turin, Turin 10154, Italy.

^6^Department of Clinical and Biological Sciences, University of Turin and S. Giovanni Bosco Hospital, Turin 10154, Italy.

**V-ATPase Consortium:**

Nancy Pinnell^1^, Dallas Reed^1^, Peter D Turnpenny^2^, Jacqueline Eason^3^, Leah Fleming^4^, Kirsty McWalter^5^, Kali Juliette^6^, Paul J Benke^7^, Xilma Ortiz-Gonzalez^8^, Sarah Mckeown^8^, Amisha B Patel^8^, Matthew Osmond^9^, Jagdeep S Walia^10^, Xianru Jiao^11^, Zhixian Yang^11^, Boris Keren^12^, Charles Perrine^12^, Ashish Deshwar^13^

^1^Floating Hospital for Children at Tufts Medical Center, Boston, MA 02111, USA.

^2^Clinical Genetics, Royal Devon & Exeter NHS Foundation Trust, Exeter EX2 5DW, UK.

^3^Nottingham Centre for Medical Genetics, Nottingham University Hospitals NHS Trust, Nottingham NG5 1PB, UK.

^4^St. Luke's Children's Hospital, Boise, Idaho 83712, USA.

^5^GeneDx, Gaithersburg, Maryland 20877, USA.

^6^Department of Neurology, Gillette Children's Specialty Healthcare, St Paul, Minnesota  55101, USA.

^7^Division of Clinical Genetics, Children's Hospital, Hollywood, Florida 33021, USA.

^8^Division of Neurology, Children’s Hospital of Philadelphia; The Epilepsy NeuroGenetics Initiative (ENGIN), Children's Hospital of Philadelphia, Pennsylvania 19104, USA.

^9^Children's Hospital of Eastern Ontario Research Institute, University of Ottawa, Ottawa K1H 8L1, Ontario, Canada.

^10^Department of Biomedical and Molecular Sciences, Queen's University, Kingston ON K7L 2V5, Canada.

^11^Department of Pediatrics, Peking University First Hospital, Beijing, China.

^12^Genetics Department, Neurogenetic Reference Center, Salpêtrière Hospital, 47 Boulevard de l'Hopital, 75013 Paris, France.

^13^Division of Clinical and Metabolic Genetics, Department of Pediatrics, The Hospital for Sick Children, University of Toronto, Toronto, Ontario M5G 1X8, Canada.
